# Maternal Controlling Parenting and Child-Perceived Mother-Child Attachment Mediate the Association between Maternal Adverse Childhood Experiences and Adolescent Behavioural Difficulties

**DOI:** 10.1101/2023.11.28.23299089

**Authors:** Yuze Shi, Shuya Xie, Angus MacBeth

## Abstract

**Background:** Maternal adverse childhood experiences (ACEs) are risk factors for increased mental health difficulties. However, the intergenerational transmission mechanisms between maternal ACEs and adolescent behavioural difficulties are poorly understood. The current study modelled the mediating effects of maternal controlling parenting and child-perceived mother-child attachment on the association between maternal ACEs and behavioural difficulties.

**Methods:** Data were obtained from Growing Up in Scotland, a nationally representative prospective probability cohort study of Scottish adolescents (*N* = 2223), followed up at 14-15 years of age. Hierarchical multivariate linear regression analyses were used to examine the strength of predictors (maternal ACEs, maternal controlling parenting, child-perceived mother-child attachment) on levels of adolescent behavioural difficulties. Mediation analyses were used to examine the serial mediating effects of maternal controlling parenting and child-perceived mother-child attachment.

**Results:** Regression analysis results indicated maternal ACEs were associated with adolescent behavioural difficulties. Serial mediation results indicated a significant indirect effect of maternal ACEs on adolescent mental health outcomes, through maternal controlling parenting and child-perceived mother-child attachment.

**Conclusion:** The study indicates that maternal controlling parenting and child-perceived mother-child attachment are mechanisms mediating the effect of maternal ACEs on adolescent behavioural difficulties. Consequently, attending to the impact of both maternal and children’s factors may improve policy programming to ameliorate the intergenerational impact of maternal ACEs on adolescents’ mental health.

**Key points:**

- Research suggests maternal adverse childhood experiences have intergenerational impacts on children.
- Maternal controlling parenting and child-perceived mother-child attachment potentially mediate the intergenerational link between maternal ACEs and behavioural difficulties of young people.
- There is a lack of research examining these mechanisms in adolescence.
- Maternal controlling parenting and child-perceived mother-child attachment mediate the link between maternal ACEs and adolescent internalizing behaviours, externalizing behaviours, and anxiety; with a small but significant serial mediating path through these two mediators.
- Policy programming should incorporate both consideration of maternal and child factors in adolescent mental health.

## Introduction

Adverse childhood experiences (ACEs), including physical, psychological, sexual abuse and neglect as well as household dysfunctions occurring prior to 18 years old (Felitti et al., 1998), are a commonly occurring public health concern affecting at least 38.8% of the global population (Hughes et al., 2021; Kessler et al., 2010). Numerous studies reports that exposure to ACEs is significantly associated with higher risk of later poor mental health conditions (Hughes et al., 2017; Panagou & MacBeth, 2022; Tzouvara et al., 2023) and may also intergenerationally adversely affect their children’s mental health (Arnold et al., 2023; Cooke et al., 2021; Moog et al., 2023). Therefore, understanding the potential mechanisms of the intergenerational transmission of parental ACEs on children’s mental health outcomes are critical for early prevention, identification, and intervention (MacBeth et al., 2023).

Alongside biological perspectives on maternal ACEs intergenerational transmission (Buss et al., 2017; Scorza et al., 2023), psychosocially informed perspective such as Bronfenbrenner’s bioecological model suggest that the multiple layers of systems around the child, form an ecological system, interacting with each other and influence child development at every life stage (Bronfenbrenner, 1977, 2001). In this type of model, the interaction between parents and the developing child is a highly influential factor, persisting into adolescence (Smetana et al., 2006).

Parenting and parent-child attachment represent two critical factors in parent-child interaction. In samples of young children, they have been identified as potential mediators of the association between maternal ACEs and child mental health outcomes (Zhang et al., 2022; Rowell & Neal-Barnett, 2022). However, it is unclear whether the effect of these mediators reduces over time or whether they continue to exert influence into adolescence. This is of particular importance given adolescence is a critical time period for biological, social, and psychological development and for the development of higher level faculties such as sense of identity and autonomy, with vulnerabilities at this stage having long term consequences for mental health and wellbeing (Blackmore, 2019). Further, research to date on mediators of the intergenerational ACEs/mental health link has focused on maternal perceptions of attachment to the child, rather than the young person’s perception of the attachment relationship.

### Potential mediators

#### Maternal Controlling Parenting

Parenting is a major component of parent-child interaction, however, growing evidence suggests mothers with ACEs are more likely to face challenges in parenting (e.g., parenting stress) and engage in negative parenting behaviours in their interactions with children (Chung et al., 2009; Madigan et al., 2019; Morris et al., 2021; Steele et al., 2016). Classically, social learning theory posits behaviours are learnt from their observations, and therefore, individuals exposed to ACEs could have learned negative parenting behaviours from their own caregivers, before intergenerationally engaging in similar behaviours in parenting their own children (Bandura, 1977; Madden et al., 2015). Equally, individuals exposed to ACEs may also develop insecure attachment characteristics, implicated in difficulties with interpersonal relationships which would further affect their capacity to provide sensitive, responsive, supportive, and consistent parenting (Bowlby, 1958, 1969; Rowell & Neal-Barnett, 2022). Reviews (Arnold et al., 2023; Rowell & Neal-Barnett, 2022; Zhang et al., 2022) suggest that although several studies have shown a significant mediating effect of maternal parenting on the association between maternal ACEs and offspring mental health outcomes (e.g., Doi,et al., 2021; Delker et al., 2014), other studies fail to observe a mediating effect (e.g., Bödeker et al., 2019; Esteves et al., 2017). Further, the majority of studies examine this the mediating effect of maternal parenting among young children or amongst samples across childhood, with few studies (Plant et al., 2017; Yoon et al., 2019) examining the mediating effect of maternal parenting among adolescents. However, adolescence is a critical stage for the development of individuals self-awareness and autonomy; therefore, parent-adolescent relationships change to accommodate these developing needs (Kobak et al., 2017; Steinberg, 2001; Morris et al., 2021). Inappropriately controlling parenting can be perceived by adolescents as overcontrolling, invasive, impinging on autonomy and may increase insecure attachment with parents and later internalizing and externalizing behaviours (e.g., Lansford et al., 2014; Soenens & Vansteenkiste, 2010, 2020). However, minimal studies (e.g., Delker et al., 2014) have examined the mediating effect of controlling parenting on the association between maternal ACEs and child psychological outcomes, and only in pre-adolescents.

#### Child-Perceived Mother-Child Attachment

The early interpersonal relationship with significant others (e.g., mother-child attachment) has a significant influence on later social, psychological, and affective functioning. Bowlby’s attachment theory posits that children’s internal working model is developed upon caregivers’ reactions and availabilities to their needs (Bowlby, 1958, 1969). If parenting traits are deemed inconsistent, unresponsive, or harsh, children may develop insecure attachment (i.e., negative internal working model) which is deemed as a risk factor for psychological and behavioural difficulties, including internalizing (Brumariu & Kerns, 2010) and externalizing behaviours (Fearon et al., 2010) and anxiety (Kerns & Brumariu, 2014). Consistent with attachment theory, research suggests negative parenting behaviours (e.g., harsh controlling) are associated with children’s insecure attachment (Fagot, 1997; Koehn & Kerns, 2018). Further, there is evidence for a mediating effect of child attachment in the association between negative parenting behaviours and poor social adjustment and externalizing behaviours of young people (Roskam et al., 2011; Xu & Yan, 2022).

However, there is still a lack of research on the relationships among maternal ACEs, mother-child attachment, and child outcomes. Only one study (Thomas-Giyer & Keesler, 2021) examined the relationship among these three constructs and found no association between maternal ACEs and mother-child attachment. However, the effect in a population participating in a family wellbeing promotion program, which may not accurately reflect the true effect as social support could buffer the influence of maternal ACEs (Thomas et al., 2018; Racine et al., 2018a; Racine et al., 2018b). Furthermore, Thomas-Giyer and Keesler (2021) used mother-perceived attachment instead of child-perceived attachment as the measure for mother-child attachment, which may underestimate the role of mother-child attachment.

#### Current Study

Although studies have evaluated the mediating effect of multiple types of parenting behaviours and mother-perceived mother-child attachment, there is a lack of research in examining the potential serial mediating effect through maternal controlling parenting and child-perceived mother-child attachment. To address these shortcomings the current study was conducted using a probability cohort sample of Scottish adolescents and their parents to examine a) the association between maternal ACEs and adolescent behavioural difficulties; and b) the serial mediating effect of maternal controlling parenting and child-perceived mother-child in the association.

## Methods

### Participants

The study used data from Growing Up in Scotland (GUS) cohort study, an ongoing longitudinal survey of a nationally representative, multistage, prospective probability sample of Scottish children (Scottish Centre for Social Research, 2022). The GUS birth cohort 1 (BC1) included 5217 10-month-old babies born between June 2004 and May 2005 and their caregivers at the first sweep in June 2005. Participated families were randomly selected using child benefit records. At sweep 9 (2017-2018), the GUS BC1 study recruited additional 502 qualified children and their families to the boost sample in adjusting the under-representation of children born to young mothers or living in the 15% most deprived areas.

We focused on the sweep 10 (2019-2020) survey at which the GUS BC1 study included 2943 cohort children and 2669 of them were from the main sample. Most cases (*N* = 2417, 82.1%) were interviewed face-to-face; however, due to COVID-19, cases which did not completed data collection through face-to-face method were invited to take part in telephone and web survey. The final sample of this study included 2223 cohort children and their families who provided valid entries in all relevant study measures and covariates, representing 75.5% of the total sample at sweep 10. Five families, which both main caregiver and partner to the caregiver were identified as mothers to the cohort children, were excluded from this study.

### Ethical Approval

Initial data collection for Growing Up in Scotland received ethical approval from the Scotland ‘A’MREC Committee (reference: 04/M RE 1 0/59). Sweep 9 and 10 received ethical approval from the Research Ethics Committee at National Centre for Social Research. The current study received ethical approval from School of Health in Social Science, The University of Edinburgh.

### Measures

#### Maternal Adverse Childhood Experiences

Maternal adverse childhood experiences (ACEs) were assessed based on the count (i.e., sum) of ten dichotomously coded ACE items occurring prior to maternal age 18: (1) parental separation, (2) household substance abuse, (3) household mental illness, (4) incarceration of a household member, (5) psychological abuse, (6) physical abuse, (7) emotional neglect, (8) physical neglect, (9) household domestic violence, (10) sexual abuse. As few individuals reported experiencing >8 ACE categories, maternal ACEs were top coded at 8 categories.

#### Maternal Controlling Parenting

Maternal controlling parenting was assessed using eight items adapted from Epstein’s Mother-Father-Peer Inventory Scale (Epstein, 1983), reported by both major caregivers and their partners at sweep 10. The measurement includes four autonomous parenting behaviour items (e.g., “I help child to become an independent person”) and four controlling parenting behaviour items (e.g., “I’m always telling child what to do and how to behave”). All items were rated on a 4-point rating scale, reverse-coded as necessary so that higher scores indicating higher controlling parenting tendency, and summed (α = 0.68 for major caregivers, α = 0.67 for partners). Scores were matched to mothers using the relationships between children and their major caregiver or partner.

#### Child-Perceived Mother-Child Attachment

Child-perceived mother-child attachment was assessed using six items adapted from People in My Life (Cook et al., 1995), which was reported by cohort children at sweep 10. All items were rated on a 4-point rating scale, and summed. Higher scores indicate more secure child-perceived mother-child attachment. At sweep 10, child-perceived mother-child attachment was collected using parent 1 and 2 method and the definition was different between face-to-face interview and telephone/web survey, further details about identifying parent 1 and 2 can be found in GUS questionnaire documentations (Scottish Centre for Social Research, 2018, 2020). These six items show high internal consistency across both parents and survey methods, ratings from 0.86 to 0.91.

#### Adolescent Behavioural Difficulties

Adolescent behavioural difficulties were assessed using the Strengths and Difficulties Questionnaire (SDQ; Goodman, 1997), administered to cohort children at sweep 10. The SDQ comprises 25 items in five equally distributed subscales: emotional symptoms, peer problems, conduct problems, hyperactivity-inattention problems, and prosocial behaviours. Emotional symptoms and peer problems subscales constitute the internalizing behaviour score; conduct problems and hyperactivity-inattention problems subscales constitute the externalizing behaviour score. All items are rated on a 3-point rating scale, reverse-coded as necessary, with higher scores indicating higher level of difficulties, summed respectively to internalizing behaviour (α = 0.78) and externalizing behaviour (α = 0.62). Goodman, Lamping, & Ploubidis (2010) recommend using the broader internalizing and externalizing behaviour scores among general population samples rather than specific subscales, due to lack of discriminant validity between individual subscales in each category.

#### Adolescent Anxiety

Adolescent anxiety was assessed with the 7-item Generalized Anxiety Disorder Assessment (Spitzer et al., 2006), reported by cohort children at sweep 10. Items were rated on a 4-point rating scale, and summed (α = 0.91). Higher scores indicate higher levels of anxiety.

#### Covariates

Considering potential confounders, the following demographic covariates were included: child’s sex (male, female, other), low birth weight (<2500g, no/yes), young motherhood at birth (under 20 year-old, no/yes), SCBU or NICU stay (no/yes), maternal ethnicity (white/other), maternal educational level (no qualification, other, lower level standard grades and vocational qualifications, upper level standard grades and intermediate vocational qualifications, higher grades and upper level vocational qualifications, degree level academic or vocational qualifications), maternal employment status (not working, working less than 16 hours per week, working more than 16 hours but less than 35 hours, working 35 hours or more), maternal risk for depression (based on three CIDI-SF qualifying questions, no/yes), maternal health and disability condition (no condition, have conditions but with no affection on daily life, have conditions with a little affection, have conditions with a lot affections), area of residence (urban/rural), social deprivation (based on Scottish Social Deprivation Index, higher score indicating less social deprivation), household equivalised income (in quintiles, higher quintile indicating higher equivalised income), family type (lone parent, couple), and number of children in household (one, two or three, four or more).

### Data Analyses

Analyses were conducted using SPSS Version 27.0 (IBM Corp., 2020). First, Pearson correlations were computed to determine associations between study measures. Second, multivariate linear regressions were performed to examine the associations between maternal ACEs and adolescent behavioural difficulties, adjusting for covariates (Model 1). Prior to conducting regression analyses, study measures were standardized. If maternal ACEs were statistically significant in Model 1, maternal controlling parenting was added into the multivariate regression models as the first potential mediator (Model 2). Subsequently, child-perceived mother-child attachment was added into the multivariate regression models as the second potential mediator (Model 3). The Process macro (Hayes, 2022) was used to examine mediating effects with 5000 times resampled bias-corrected bootstrapping used to generate 95% confidence intervals; with confidence intervals omitting zero, indicative of a statistically significant mediating effect (Hayes, 2009). All significance levels were set at p<0.05.

## Results

### Preliminary analyses

There were no significant differences between the analysed sample and the full GUS sample (Table 1). Demographic characteristics for the analysed sample are presented in Table 2. Table 3 shows Pearson correlations between study measures with Maternal ACEs, maternal controlling parenting, and child-perceived mother-child attachment all significantly correlated with adolescent internalizing, externalizing behaviours, and anxiety. Maternal ACEs were significantly correlated with maternal controlling parenting and child-perceived mother-child attachment. Maternal controlling parenting was also significantly correlated with child-perceived mother-child attachment.

**Table 1.** Comparison of study measures between full and analysing sample.

| Variables | Full sample |  |  | Analysing sample |  |  |
| --- | --- | --- | --- | --- | --- | --- |
|  | *M* | *SD* | *N* | *M* | *SD* | *N* |
| Maternal ACEs | 2.45 | 2.21 | 2633 | 2.46 | 2.21 | 2223 |
| Maternal controlling parenting | 14.12 | 3.49 | 2638 | 14.00 | 3.42 | 2223 |
| Child-perceived mother-child attachment | 20.33 | 3.79 | 2554 | 20.31 | 3.82 | 2223 |
| Adolescent internalizing behaviors | 5.24 | 3.68 | 2621 | 5.23 | 3.69 | 2223 |
| Adolescent externalizing behaviors | 6.13 | 2.90 | 2621 | 6.13 | 2.91 | 2223 |
| Adolescent anxiety | 4.68 | 4.93 | 2629 | 4.73 | 4.96 | 2223 |

**Table 2.**
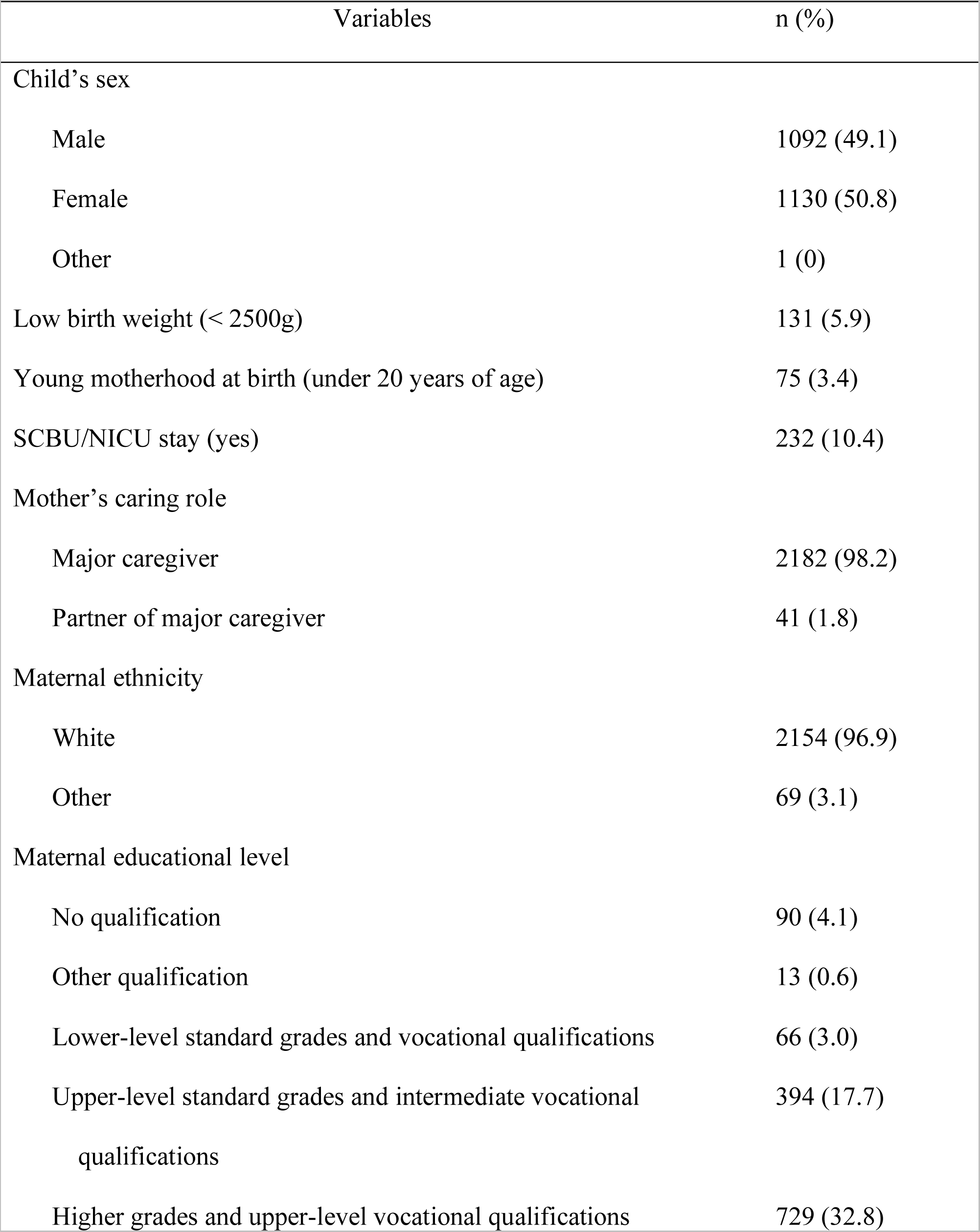
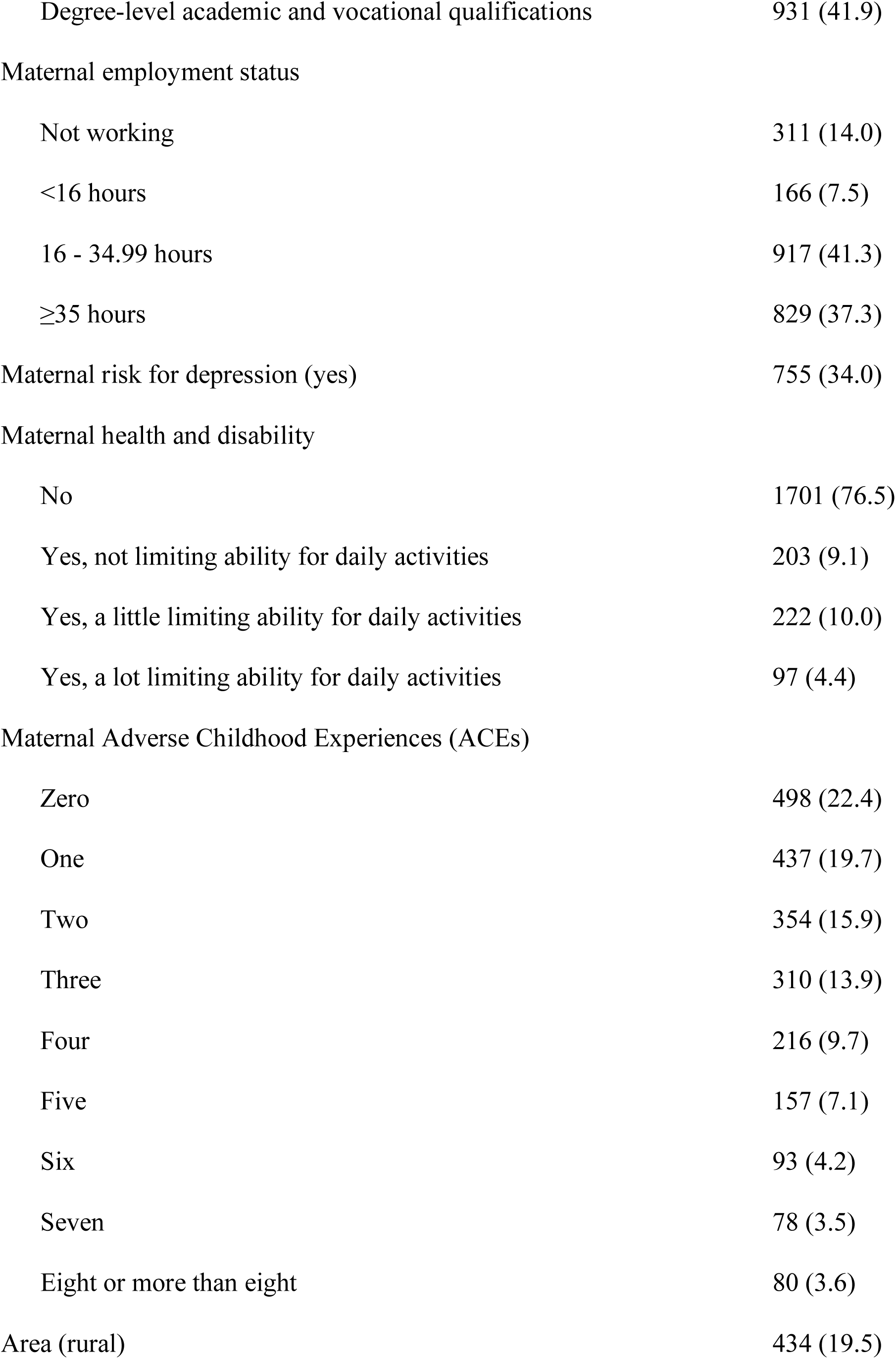
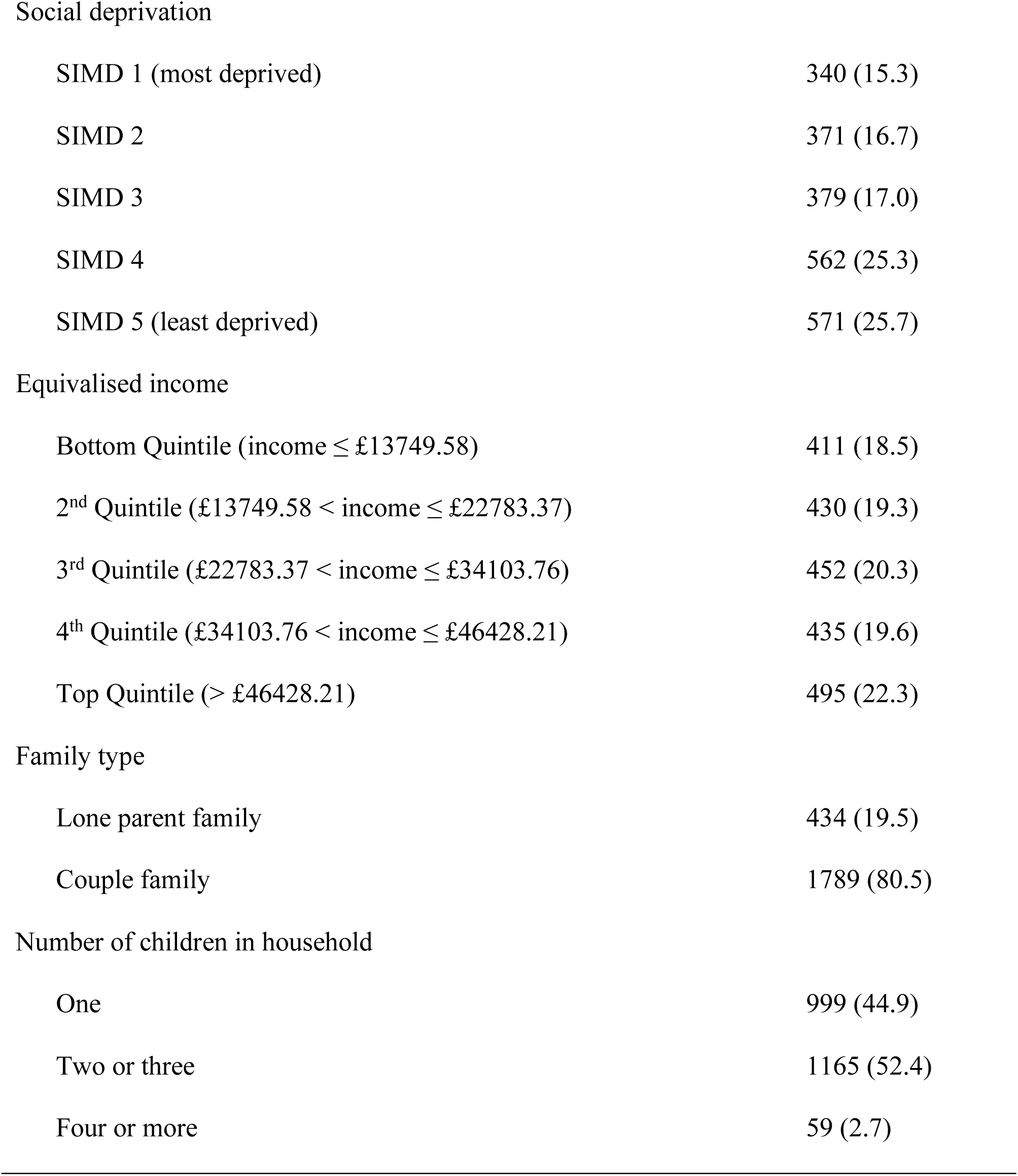
Demographic information of the participants in the study (N = 2223)

| Variables | n (%) |
| --- | --- |
| Child's sex |  |
| Male | 1092 (49.1) |
| Female | 1130 (50.8) |
| Other | 1 (0) |
| Low birth weight (< 2500g) | 131 (5.9) |
| Young motherhood at birth (under 20 years of age) | 75 (3.4) |
| SCBU/NICU stay (yes) | 232 (10.4) |
| Mother's caring role |  |
| Major caregiver | 2182 (98.2) |
| Partner of major caregiver | 41 (1.8) |
| Maternal ethnicity |  |
| White | 2154 (96.9) |
| Other | 69 (3.1) |
| Maternal educational level |  |
| No qualification | 90 (4.1) |
| Other qualification | 13 (0.6) |
| Lower-level standard grades and vocational qualifications | 66 (3.0) |
| Upper-level standard grades and intermediate vocational qualifications | 394 (17.7) |
| Higher grades and upper-level vocational qualifications | 729 (32.8) |
| Degree-level academic and vocational qualifications | 931 (41.9) |
| Maternal employment status |  |
| Not working | 311 (14.0) |
| <16 hours | 166 (7.5) |
| 16 - 34.99 hours | 917 (41.3) |
| ≥35 hours | 829 (37.3) |
| Maternal risk for depression (yes) | 755 (34.0) |
| Maternal health and disability |  |
| No | 1701 (76.5) |
| Yes, not limiting ability for daily activities | 203 (9.1) |
| Yes, a little limiting ability for daily activities | 222 (10.0) |
| Yes, a lot limiting ability for daily activities | 97 (4.4) |
| Maternal Adverse Childhood Experiences (ACEs) |  |
| Zero | 498 (22.4) |
| One | 437 (19.7) |
| Two | 354 (15.9) |
| Three | 310 (13.9) |
| Four | 216 (9.7) |
| Five | 157 (7.1) |
| Six | 93 (4.2) |
| Seven | 78 (3.5) |
| Eight or more than eight | 80 (3.6) |
| Area (rural) | 434 (19.5) |
| Social deprivation |  |
| SIMD 1 (most deprived) | 340 (15.3) |
| SIMD 2 | 371 (16.7) |
| SIMD 3 | 379 (17.0) |
| SIMD 4 | 562 (25.3) |
| SIMD 5 (least deprived) | 571 (25.7) |
| Equivalised income |  |
| Bottom Quintile (income ≤ £13749.58) | 411 (18.5) |
| 2<sup>nd</sup> Quintile (£13749.58 < income ≤ £22783.37) | 430 (19.3) |
| 3<sup>rd</sup> Quintile (£22783.37 < income ≤ £34103.76) | 452 (20.3) |
| 4<sup>th</sup> Quintile (£34103.76 < income ≤ £46428.21) | 435 (19.6) |
| Top Quintile (> £46428.21) | 495 (22.3) |
| Family type |  |
| Lone parent family | 434 (19.5) |
| Couple family | 1789 (80.5) |
| Number of children in household |  |
| One | 999 (44.9) |
| Two or three | 1165 (52.4) |
| Four or more | 59 (2.7) |

**Table 3.** Pearson’s correlations for study measures.

| Variable | 1 | 2 | 3 | 4 | 5 | 6 |
| --- | --- | --- | --- | --- | --- | --- |
| 1. Maternal ACEs | — |  |  |  |  |  |
| 2. Maternal controlling parenting | .16*** | — |  |  |  |  |
| 3. Child-perceived mother-child attachment | -.12*** | -.11*** | — |  |  |  |
| 4. Adolescent Internalizing Behaviours | .15*** | .10*** | -.31*** | — |  |  |
| 5. Adolescent Externalizing Behaviours | .12*** | .21*** | -.36*** | .45*** | — |  |
| 6. Adolescent Anxiety | .13*** | .08*** | -.34*** | .70*** | .50*** | — |

Note. ACEs: Adverse Childhood Experience. \*\*\* $p < .001$ . $N = 2223$ .

### Regression analyses

In the hierarchical multivariate linear regression analyses (Table 4), after adjusting for covariates in the model, maternal ACEs were significantly associated with adolescent internalizing and externalizing behaviours, and anxiety (model 1). After adding maternal controlling parenting into regression analyses as the first potential mediator (model 2), both maternal ACEs and maternal controlling parenting were each significantly associated with all three adolescent behavioural outcomes. After further adding child-perceived mother-child attachment into regression analyses as the second potential mediator (model 3), maternal ACEs, maternal controlling parenting, and child-perceived mother-child attachment were each significantly associated with adolescent internalizing behaviours and anxiety. However, maternal controlling parenting, and child-perceived mother-child attachment, but not maternal ACEs, were each significantly associated with adolescent externalizing behaviours in model 3, suggesting the association between maternal ACEs and adolescent externalizing behaviours might be completely mediated by the two factors.

**Table 4.**
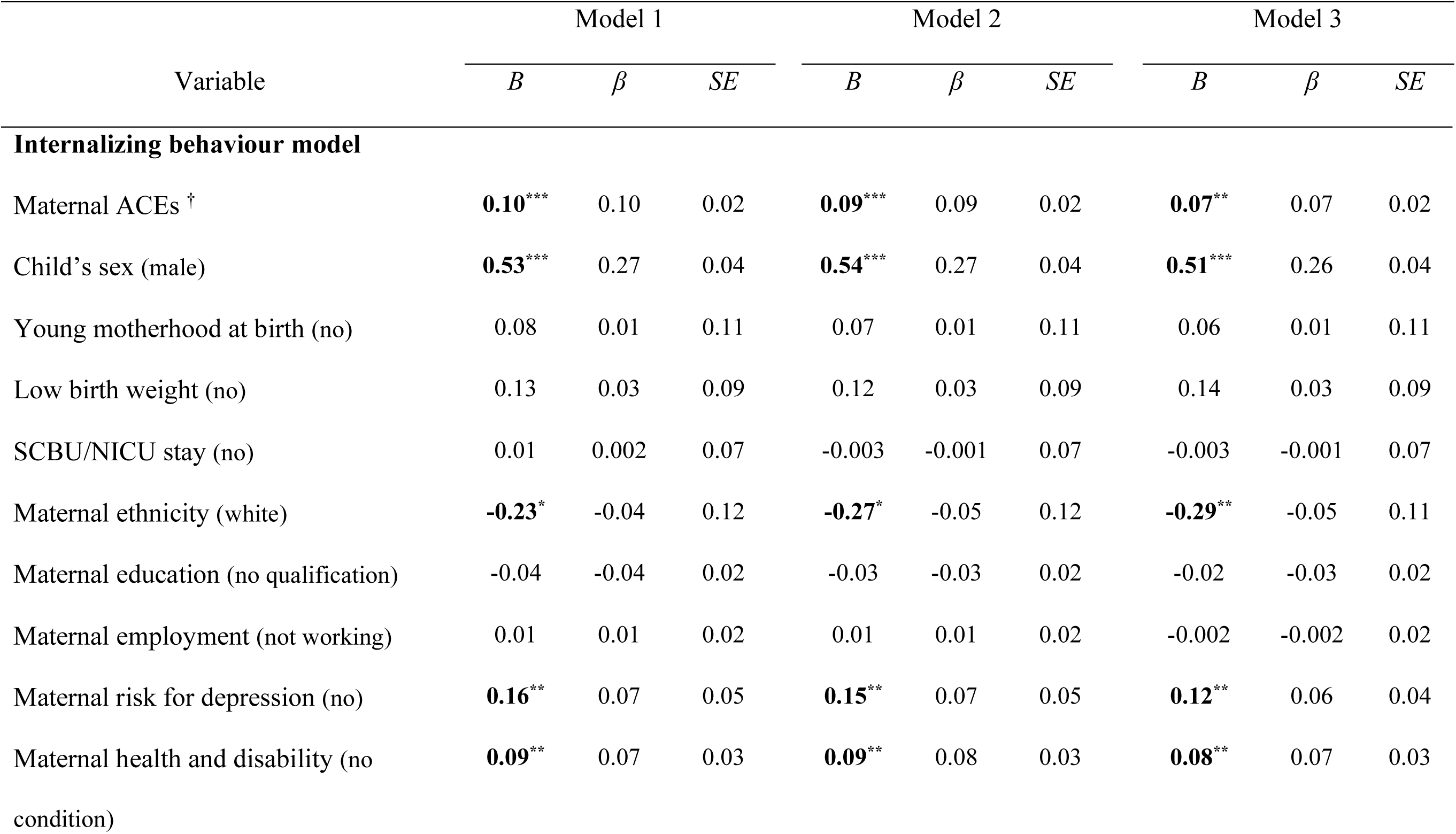
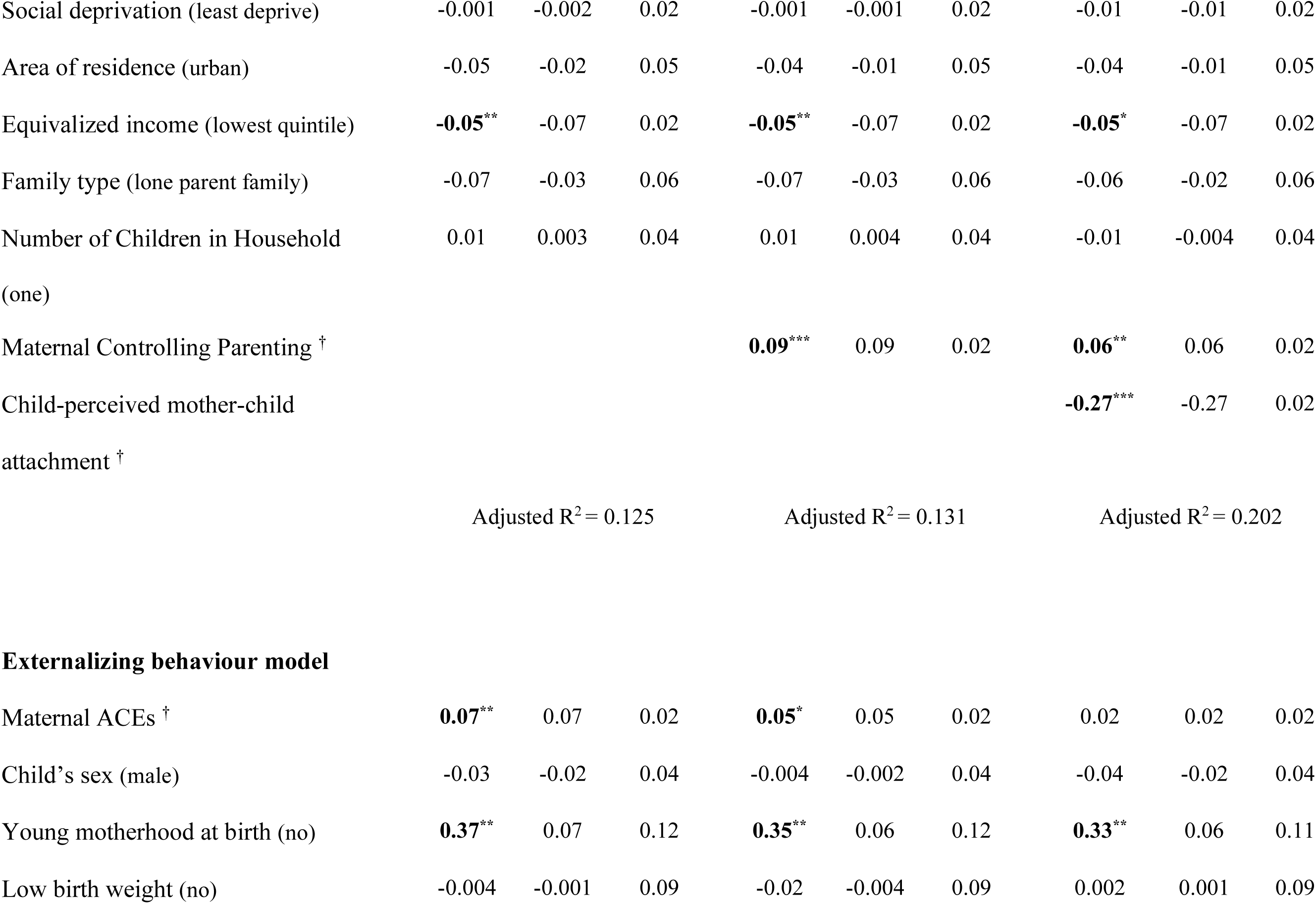
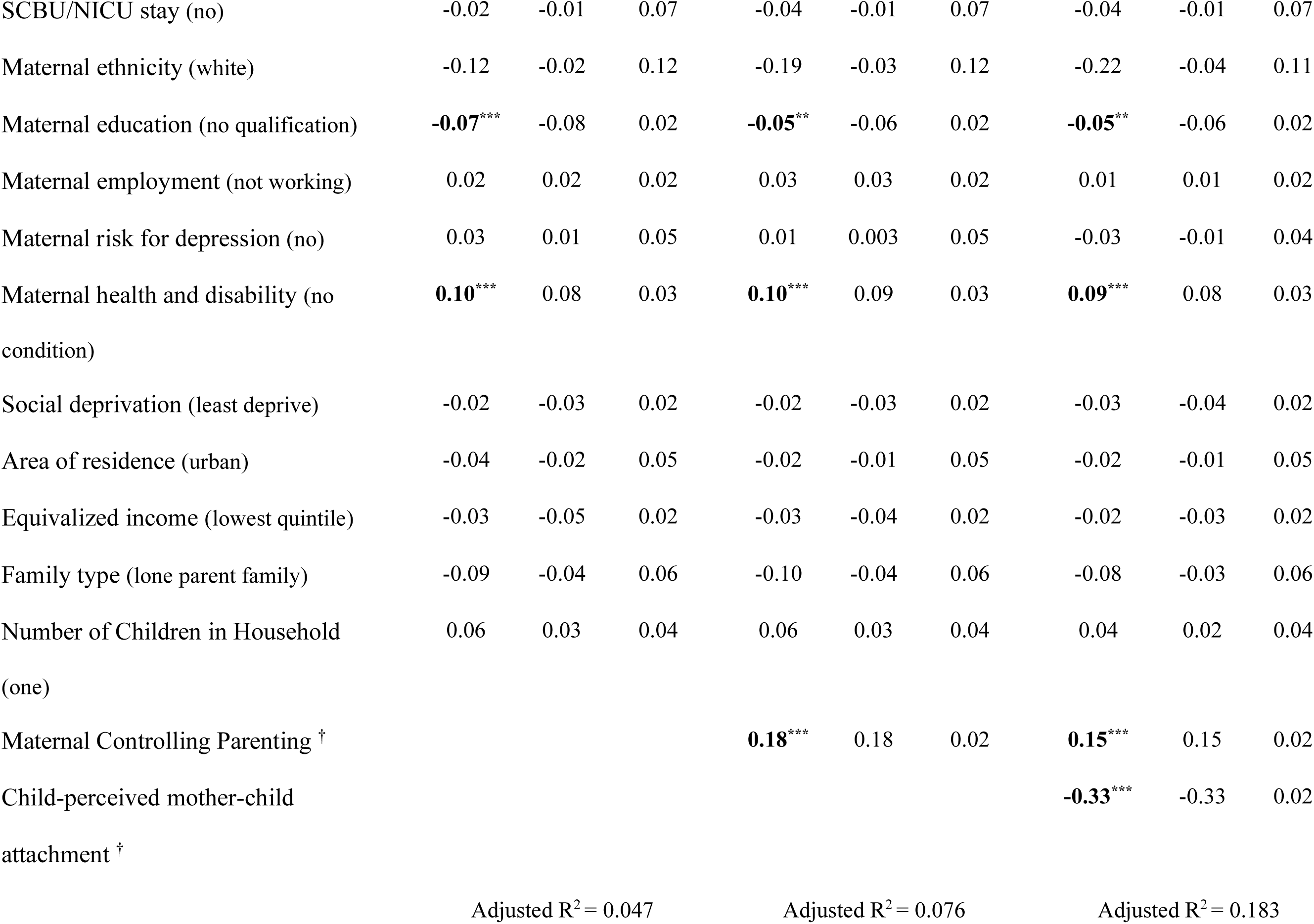
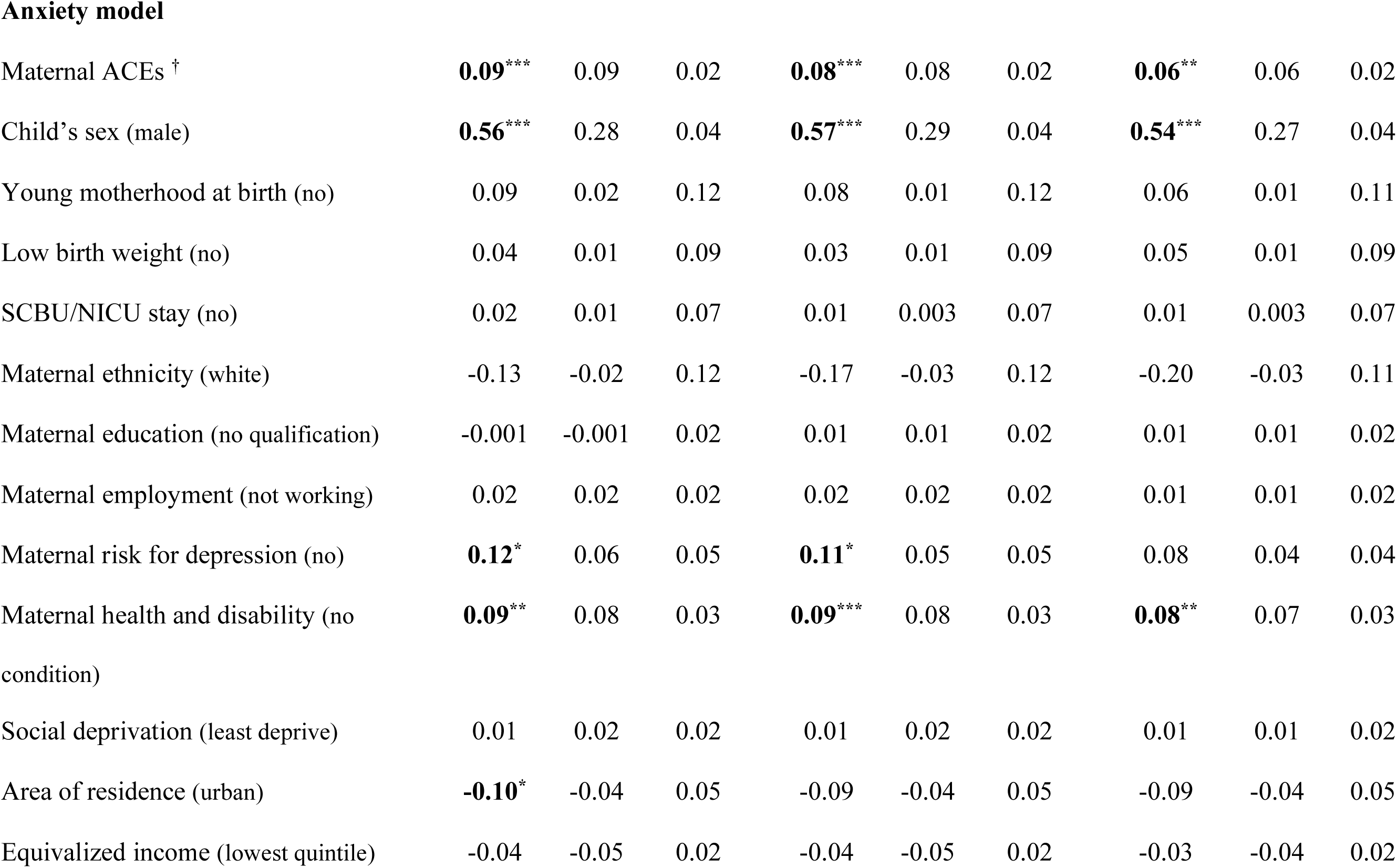
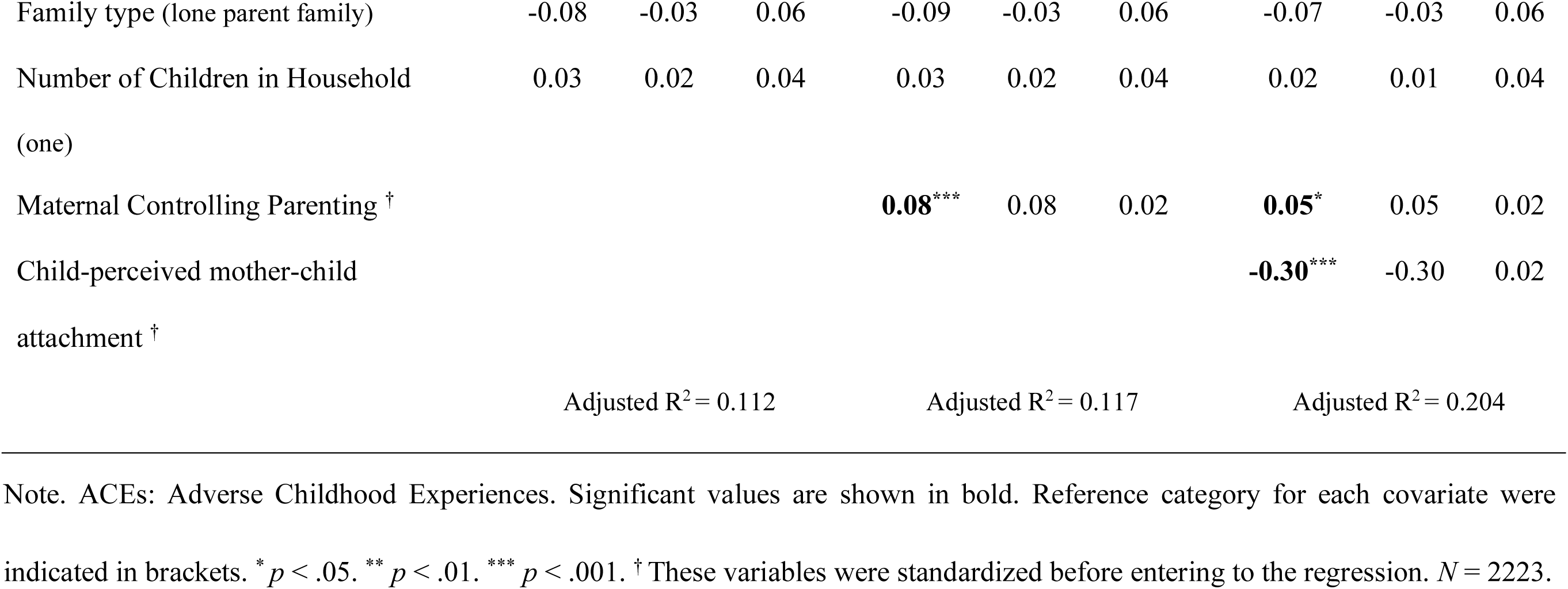
Results of hierarchical regression analyses on adolescent behavioural difficulties.

| Variable | Model 1 |  |  | Model 2 |  |  | Model 3 |  |  |
| --- | --- | --- | --- | --- | --- | --- | --- | --- | --- |
| | *B* | $\beta$ | *SE* | *B* | $\beta$ | *SE* | *B* | $\beta$ | *SE* |
| **Internalizing behaviour model** |  |  |  |  |  |  |  |  |  |
| Maternal ACEs <sup>†</sup> | **0.10\*\*\*** | 0.10 | 0.02 | **0.09\*\*\*** | 0.09 | 0.02 | **0.07\*\*** | 0.07 | 0.02 |
| Child's sex (male) | **0.53\*\*\*** | 0.27 | 0.04 | **0.54\*\*\*** | 0.27 | 0.04 | **0.51\*\*\*** | 0.26 | 0.04 |
| Young motherhood at birth (no) | 0.08 | 0.01 | 0.11 | 0.07 | 0.01 | 0.11 | 0.06 | 0.01 | 0.11 |
| Low birth weight (no) | 0.13 | 0.03 | 0.09 | 0.12 | 0.03 | 0.09 | 0.14 | 0.03 | 0.09 |
| SCBU/NICU stay (no) | 0.01 | 0.002 | 0.07 | -0.003 | -0.001 | 0.07 | -0.003 | -0.001 | 0.07 |
| Maternal ethnicity (white) | **-0.23\*** | -0.04 | 0.12 | **-0.27\*** | -0.05 | 0.12 | **-0.29\*\*** | -0.05 | 0.11 |
| Maternal education (no qualification) | -0.04 | -0.04 | 0.02 | -0.03 | -0.03 | 0.02 | -0.02 | -0.03 | 0.02 |
| Maternal employment (not working) | 0.01 | 0.01 | 0.02 | 0.01 | 0.01 | 0.02 | -0.002 | -0.002 | 0.02 |
| Maternal risk for depression (no) | **0.16\*\*** | 0.07 | 0.05 | **0.15\*\*** | 0.07 | 0.05 | **0.12\*\*** | 0.06 | 0.04 |
| Maternal health and disability (no condition) | **0.09\*\*** | 0.07 | 0.03 | **0.09\*\*** | 0.08 | 0.03 | **0.08\*\*** | 0.07 | 0.03 |
| Social deprivation (least deprive) | -0.001 | -0.002 | 0.02 | -0.001 | -0.001 | 0.02 | -0.01 | -0.01 | 0.02 |
| Area of residence (urban) | -0.05 | -0.02 | 0.05 | -0.04 | -0.01 | 0.05 | -0.04 | -0.01 | 0.05 |
| Equivalized income (lowest quintile) | **-0.05\*\*** | -0.07 | 0.02 | **-0.05\*\*** | -0.07 | 0.02 | **-0.05\*** | -0.07 | 0.02 |
| Family type (lone parent family) | -0.07 | -0.03 | 0.06 | -0.07 | -0.03 | 0.06 | -0.06 | -0.02 | 0.06 |
| Number of Children in Household<br>(one) | 0.01 | 0.003 | 0.04 | 0.01 | 0.004 | 0.04 | -0.01 | -0.004 | 0.04 |
| Maternal Controlling Parenting <sup>†</sup> |  |  |  | **0.09\*\*\*** | 0.09 | 0.02 | **0.06\*\*** | 0.06 | 0.02 |
| Child-perceived mother-child<br>attachment <sup>†</sup> |  |  |  |  |  |  | **-0.27\*\*\*** | -0.27 | 0.02 |
|  | Adjusted R<sup>2</sup> = 0.125 |  |  | Adjusted R<sup>2</sup> = 0.131 |  |  | Adjusted R<sup>2</sup> = 0.202 |  |  |
| Externalizing behaviour model |  |  |  |  |  |  |  |  |  |
| Maternal ACEs <sup>†</sup> | **0.07\*\*** | 0.07 | 0.02 | **0.05\*** | 0.05 | 0.02 | 0.02 | 0.02 | 0.02 |
| Child's sex (male) | -0.03 | -0.02 | 0.04 | -0.004 | -0.002 | 0.04 | -0.04 | -0.02 | 0.04 |
| Young motherhood at birth (no) | **0.37\*\*** | 0.07 | 0.12 | **0.35\*\*** | 0.06 | 0.12 | **0.33\*\*** | 0.06 | 0.11 |
| Low birth weight (no) | -0.004 | -0.001 | 0.09 | -0.02 | -0.004 | 0.09 | 0.002 | 0.001 | 0.09 |
| SCBU/NICU stay (no) | -0.02 | -0.01 | 0.07 | -0.04 | -0.01 | 0.07 | -0.04 | -0.01 | 0.07 |
| Maternal ethnicity (white) | -0.12 | -0.02 | 0.12 | -0.19 | -0.03 | 0.12 | -0.22 | -0.04 | 0.11 |
| Maternal education (no qualification) | **-0.07\*\*\*** | -0.08 | 0.02 | **-0.05\*\*** | -0.06 | 0.02 | **-0.05\*\*** | -0.06 | 0.02 |
| Maternal employment (not working) | 0.02 | 0.02 | 0.02 | 0.03 | 0.03 | 0.02 | 0.01 | 0.01 | 0.02 |
| Maternal risk for depression (no) | 0.03 | 0.01 | 0.05 | 0.01 | 0.003 | 0.05 | -0.03 | -0.01 | 0.04 |
| Maternal health and disability (no condition) | **0.10\*\*\*** | 0.08 | 0.03 | **0.10\*\*\*** | 0.09 | 0.03 | **0.09\*\*\*** | 0.08 | 0.03 |
| Social deprivation (least deprive) | -0.02 | -0.03 | 0.02 | -0.02 | -0.03 | 0.02 | -0.03 | -0.04 | 0.02 |
| Area of residence (urban) | -0.04 | -0.02 | 0.05 | -0.02 | -0.01 | 0.05 | -0.02 | -0.01 | 0.05 |
| Equivalized income (lowest quintile) | -0.03 | -0.05 | 0.02 | -0.03 | -0.04 | 0.02 | -0.02 | -0.03 | 0.02 |
| Family type (lone parent family) | -0.09 | -0.04 | 0.06 | -0.10 | -0.04 | 0.06 | -0.08 | -0.03 | 0.06 |
| Number of Children in Household (one) | 0.06 | 0.03 | 0.04 | 0.06 | 0.03 | 0.04 | 0.04 | 0.02 | 0.04 |
| Maternal Controlling Parenting <sup>†</sup> |  |  |  | **0.18\*\*\*** | 0.18 | 0.02 | **0.15\*\*\*** | 0.15 | 0.02 |
| Child-perceived mother-child attachment <sup>†</sup> |  |  |  |  |  |  | **-0.33\*\*\*** | -0.33 | 0.02 |
|  | Adjusted R<sup>2</sup> = 0.047 |  |  | Adjusted R<sup>2</sup> = 0.076 |  |  | Adjusted R<sup>2</sup> = 0.183 |  |  |
| **Anxiety model** |  |  |  |  |  |  |  |  |  |
| Maternal ACEs <sup>†</sup> | **0.09\*\*\*** | 0.09 | 0.02 | **0.08\*\*\*** | 0.08 | 0.02 | **0.06\*\*** | 0.06 | 0.02 |
| Child's sex (male) | **0.56\*\*\*** | 0.28 | 0.04 | **0.57\*\*\*** | 0.29 | 0.04 | **0.54\*\*\*** | 0.27 | 0.04 |
| Young motherhood at birth (no) | 0.09 | 0.02 | 0.12 | 0.08 | 0.01 | 0.12 | 0.06 | 0.01 | 0.11 |
| Low birth weight (no) | 0.04 | 0.01 | 0.09 | 0.03 | 0.01 | 0.09 | 0.05 | 0.01 | 0.09 |
| SCBU/NICU stay (no) | 0.02 | 0.01 | 0.07 | 0.01 | 0.003 | 0.07 | 0.01 | 0.003 | 0.07 |
| Maternal ethnicity (white) | -0.13 | -0.02 | 0.12 | -0.17 | -0.03 | 0.12 | -0.20 | -0.03 | 0.11 |
| Maternal education (no qualification) | -0.001 | -0.001 | 0.02 | 0.01 | 0.01 | 0.02 | 0.01 | 0.01 | 0.02 |
| Maternal employment (not working) | 0.02 | 0.02 | 0.02 | 0.02 | 0.02 | 0.02 | 0.01 | 0.01 | 0.02 |
| Maternal risk for depression (no) | **0.12\*** | 0.06 | 0.05 | **0.11\*** | 0.05 | 0.05 | 0.08 | 0.04 | 0.04 |
| Maternal health and disability (no condition) | **0.09\*\*** | 0.08 | 0.03 | **0.09\*\*\*** | 0.08 | 0.03 | **0.08\*\*** | 0.07 | 0.03 |
| Social deprivation (least deprive) | 0.01 | 0.02 | 0.02 | 0.01 | 0.02 | 0.02 | 0.01 | 0.01 | 0.02 |
| Area of residence (urban) | **-0.10\*** | -0.04 | 0.05 | -0.09 | -0.04 | 0.05 | -0.09 | -0.04 | 0.05 |
| Equalized income (lowest quintile) | -0.04 | -0.05 | 0.02 | -0.04 | -0.05 | 0.02 | -0.03 | -0.04 | 0.02 |
| Family type (lone parent family) | -0.08 | -0.03 | 0.06 | -0.09 | -0.03 | 0.06 | -0.07 | -0.03 | 0.06 |
| Number of Children in Household<br>(one) | 0.03 | 0.02 | 0.04 | 0.03 | 0.02 | 0.04 | 0.02 | 0.01 | 0.04 |
| Maternal Controlling Parenting <sup>†</sup> |  |  |  | **0.08\*\*\*** | 0.08 | 0.02 | **0.05\*** | 0.05 | 0.02 |
| Child-perceived mother-child<br>attachment <sup>†</sup> |  |  |  |  |  |  | **-0.30\*\*\*** | -0.30 | 0.02 |
|  | Adjusted R<sup>2</sup> = 0.112 |  |  | Adjusted R<sup>2</sup> = 0.117 |  |  | Adjusted R<sup>2</sup> = 0.204 |  |  |

Note. ACEs: Adverse Childhood Experiences. Significant values are shown in bold. Reference category for each covariate were indicated in brackets. \* $p < .05$ . \*\* $p < .01$ . \*\*\* $p < .001$ . <sup>†</sup> These variables were standardized before entering to the regression. $N = 2223$ .

### Mediation analyses

Next, serial mediation analyses were conducted to further examine whether maternal controlling parenting and child-perceived parent-child attachment mediate the association between maternal ACEs and adolescent mental health outcomes. Serial mediation models and 95% confidence interval bootstrapping results are presented in Figure 1-3 and Table 5 respectively.

**Figure 1.**
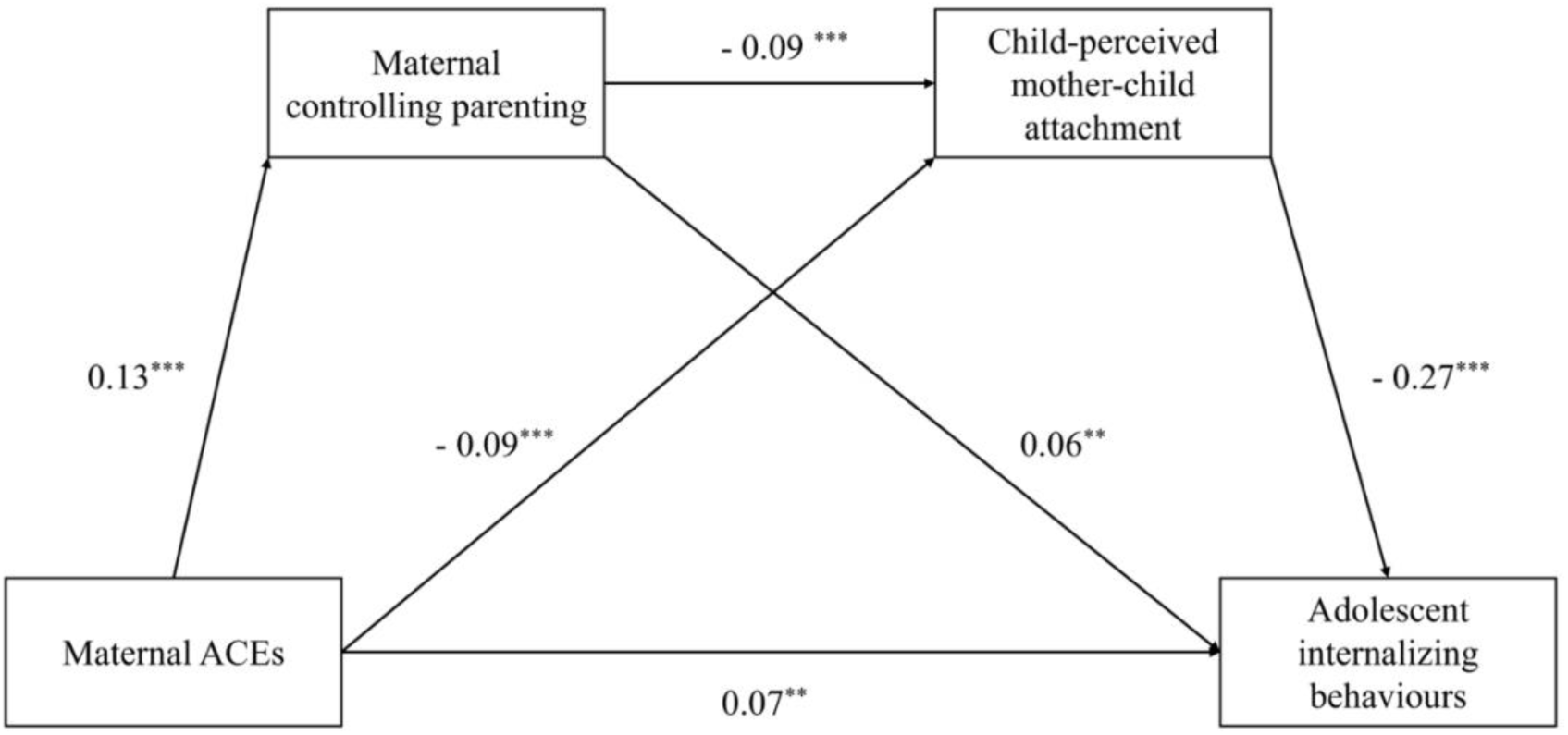
Serial mediation model on adolescent internalizing behaviours. Note. Standardized coefficients are reported. Controlled covariates. ** *p* < 0.01; *** *p* < 0.001.

**Table 5.**
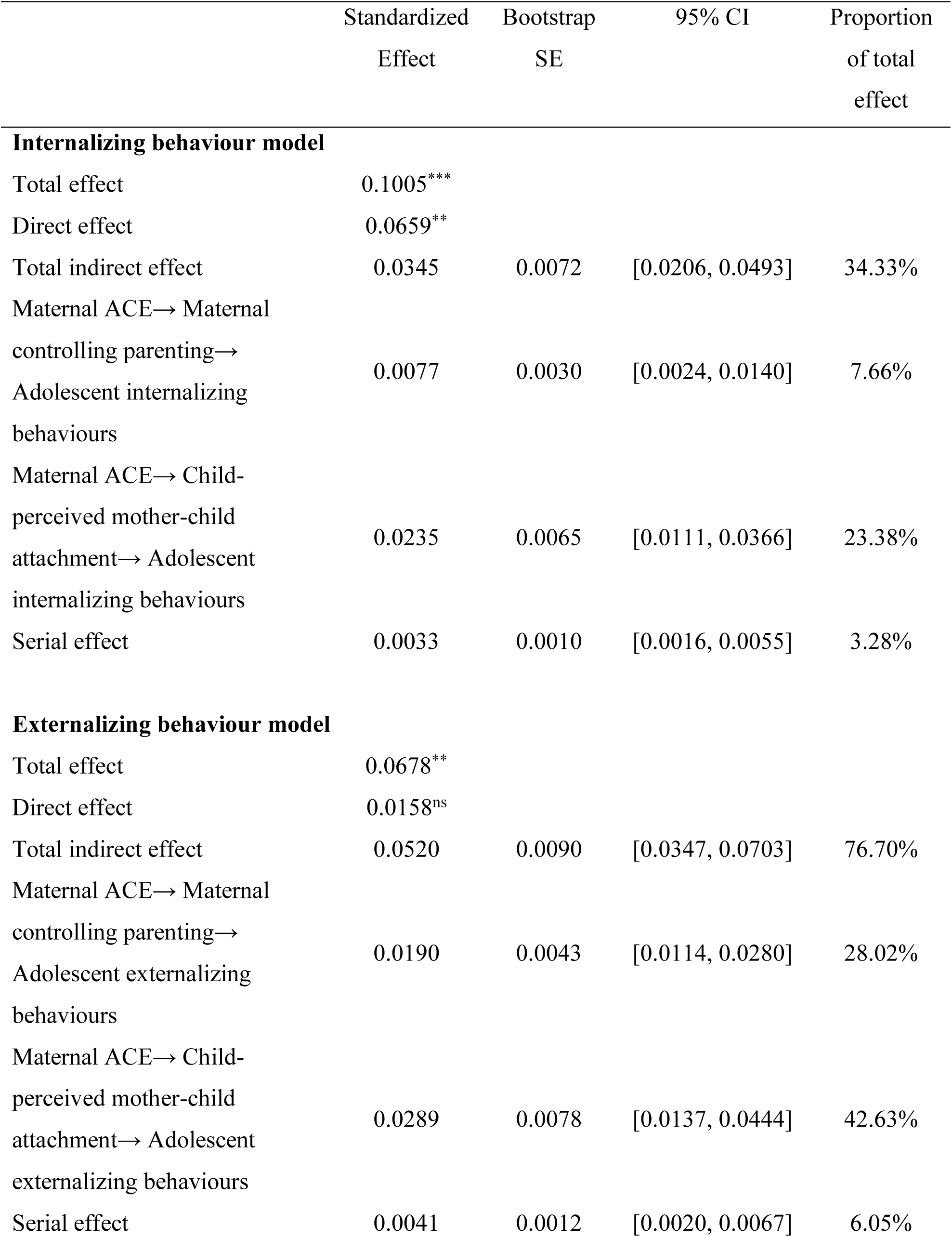
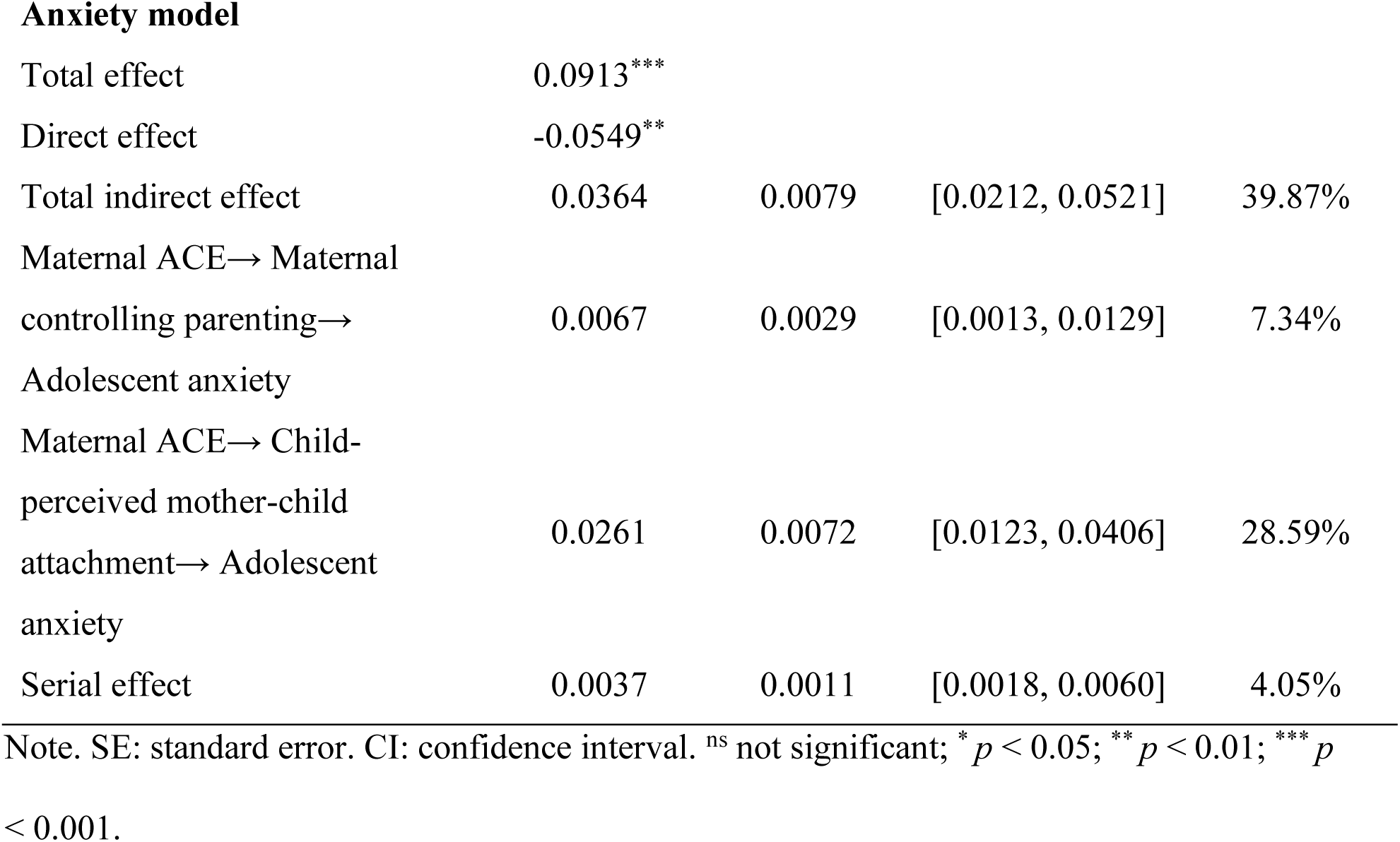
Results of serial mediation analyses on adolescent behavioural difficulties.

|  | Standardized<br>Effect | Bootstrap<br>SE | 95% CI | Proportion<br>of total<br>effect |
| --- | --- | --- | --- | --- |
| **Internalizing behaviour model** |  |  |  |  |
| Total effect | 0.1005*** |  |  |  |
| Direct effect | 0.0659** |  |  |  |
| Total indirect effect | 0.0345 | 0.0072 | [0.0206, 0.0493] | 34.33% |
| Maternal ACE→ Maternal<br>controlling parenting→<br>Adolescent internalizing<br>behaviours | 0.0077 | 0.0030 | [0.0024, 0.0140] | 7.66% |
| Maternal ACE→ Child-perceived mother-child<br>attachment→ Adolescent<br>internalizing behaviours | 0.0235 | 0.0065 | [0.0111, 0.0366] | 23.38% |
| Serial effect | 0.0033 | 0.0010 | [0.0016, 0.0055] | 3.28% |
| **Externalizing behaviour model** |  |  |  |  |
| Total effect | 0.0678** |  |  |  |
| Direct effect | 0.0158<sup>ns</sup> |  |  |  |
| Total indirect effect | 0.0520 | 0.0090 | [0.0347, 0.0703] | 76.70% |
| Maternal ACE→ Maternal<br>controlling parenting→<br>Adolescent externalizing<br>behaviours | 0.0190 | 0.0043 | [0.0114, 0.0280] | 28.02% |
| Maternal ACE→ Child-perceived mother-child<br>attachment→ Adolescent<br>externalizing behaviours | 0.0289 | 0.0078 | [0.0137, 0.0444] | 42.63% |
| Serial effect | 0.0041 | 0.0012 | [0.0020, 0.0067] | 6.05% |
| **Anxiety model** |  |  |  |  |
| Total effect | 0.0913*** |  |  |  |
| Direct effect | -0.0549** |  |  |  |
| Total indirect effect | 0.0364 | 0.0079 | [0.0212, 0.0521] | 39.87% |
| Maternal ACE→ Maternal<br>controlling parenting→<br>Adolescent anxiety | 0.0067 | 0.0029 | [0.0013, 0.0129] | 7.34% |
| Maternal ACE→ Child-perceived mother-child<br>attachment→ Adolescent<br>anxiety | 0.0261 | 0.0072 | [0.0123, 0.0406] | 28.59% |
| Serial effect | 0.0037 | 0.0011 | [0.0018, 0.0060] | 4.05% |

Note. SE: standard error. CI: confidence interval. <sup>ns</sup> not significant; \* $p < 0.05$ ; \*\* $p < 0.01$ ; \*\*\* $p < 0.001$ .

Figure 1 present the serial mediation model for adolescent internalizing behaviours. Results indicated a significant indirect effect of maternal ACEs on adolescent internalizing behaviours through maternal controlling parenting and child-perceived mother-child attachment, explaining 21% of the variance in adolescent internalizing behaviours. There were also significant indirect effects through maternal controlling parenting and child-perceived mother-child attachment individually.

Results for adolescent externalizing behaviours (Figure 2) indicated a significant indirect effect of maternal ACEs on adolescent externalizing behaviours through maternal controlling parenting and child-perceived mother-child attachment. There were also significant indirect effects through maternal controlling parenting and child-perceived mother-child attachment individually. The serial mediation model explained 19% of the variance of adolescent externalizing behaviours. Notably, the direct effect was non-significant in this model which suggests maternal controlling parenting and child-perceived mother-child attachment completely mediated the association between maternal ACEs and adolescent externalizing behaviours.

**Figure 2.**
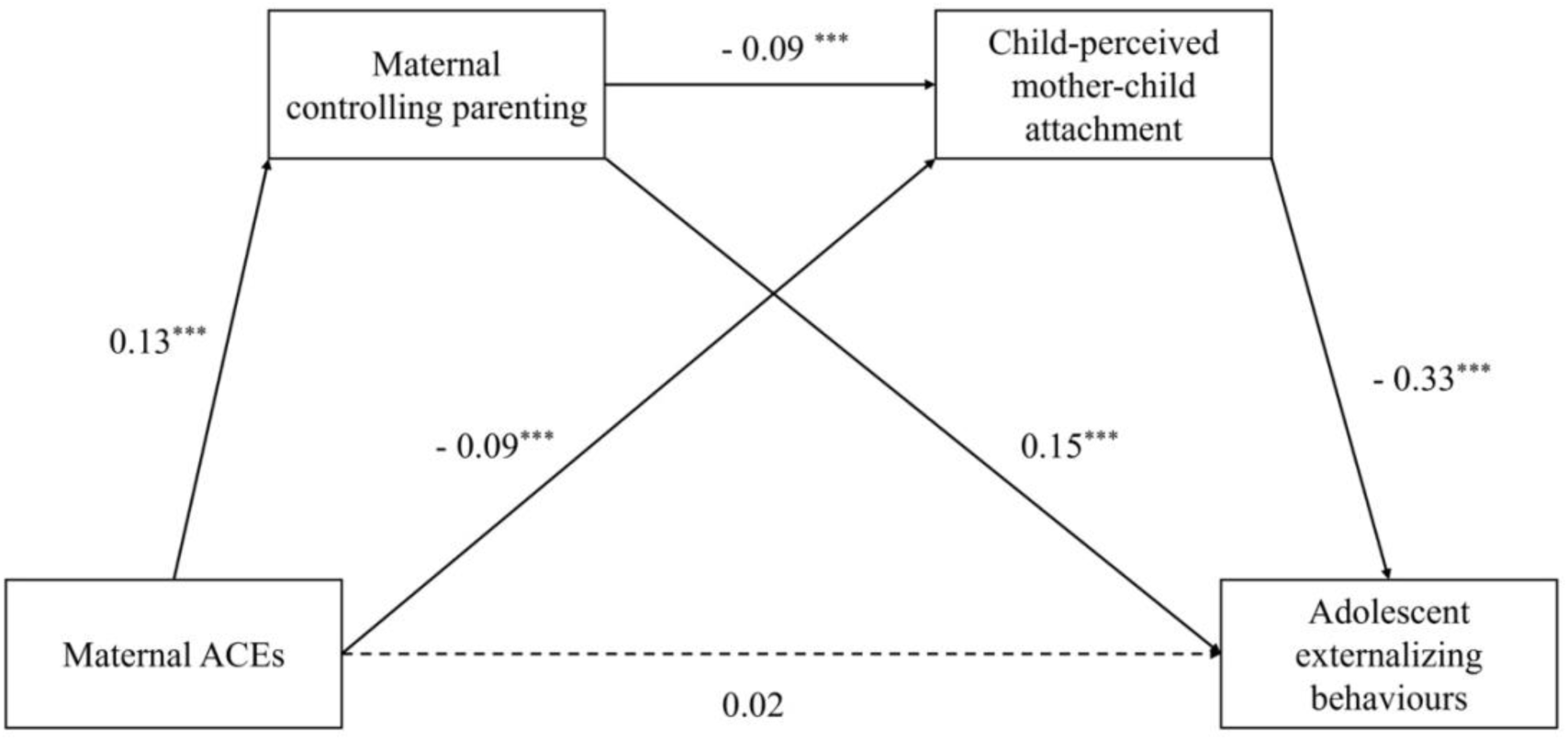
Serial mediation model on adolescent externalizing behaviours. Note. Standardized coefficients are reported. Controlled covariates. *** *p* < 0.001. Non-significant path is shown in dotted line, *p* = .423.

The serial mediation analysis for adolescent anxiety (Figure 3) indicated a significant indirect effect of maternal ACEs on adolescent anxiety through maternal controlling parenting and child-perceived mother-child attachment. There were also significant indirect effects through maternal controlling parenting and child-perceived mother-child attachment individually. The serial mediation model explained 21% of the variance of adolescent anxiety. Results in the serial mediation analyses also indicated maternal controlling parenting were also affected by child sex (*β* = -0.08, *p* < 0.001), maternal ethnicity (*β* = 0.07, *p* = 0.001), maternal education level (*β* = -0.11, *p* < 0.001), maternal employment status (*β* = -0.06, *p* = 0.006), maternal risk for depression (*β* = 0.05, *p* = 0.034), and area of residence (i.e., urban/rural, *β* = -0.04, *p* = 0.041). Current serial mediation analyses also found, when controlling maternal ACEs and maternal controlling parenting, child sex (*β* = -0.06, *p* = 0.005), maternal employment status (*β* = -0.05, *p* = 0.032) and maternal risk for depression (*β* = -0.05, *p* = 0.038) significantly predicted child-perceived mother child attachment.

**Figure 3.**
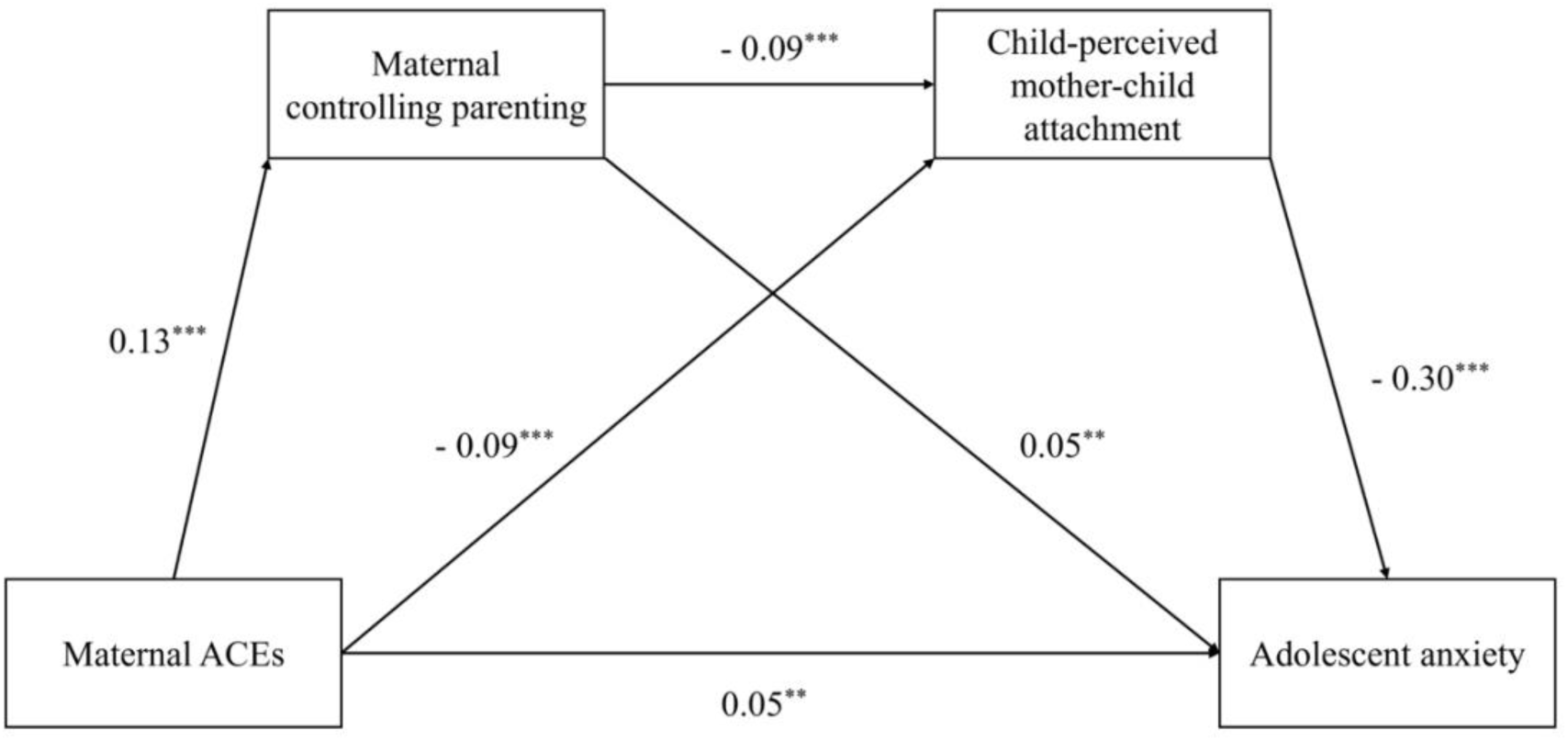
Serial mediation model on adolescent anxiety. Note. Standardized coefficients are reported. Controlled covariates. * *p* < 0.05; ** *p* < 0.01; *** *p* < 0.001.

## Discussion

This study examined whether maternal controlling parenting and child-perceived mother-child attachment mediate the association between maternal ACEs and adolescent behavioural difficulties. To the authors’ knowledge, this study is the first to examine mediating effects of both maternal controlling parenting and child-perceived mother-child attachment on the above outcomes in an adolescent sample.

Supporting our hypotheses, maternal ACEs were positively associated with adolescent internalizing, externalizing behaviours, and anxiety. This is consistent with the previous literature, reporting robust intergenerational associations between maternal ACEs and mental health outcomes of children and adolescents (e.g., Arnold et al., 2023; Clayborne et al., 2021; Cooke et al., 2021).

Furthermore, we found a significant indirect effect via maternal controlling parenting and child-perceived mother-child attachment in the association between maternal ACEs and all four adolescent behavioural outcomes. This study also found significant indirect effects through controlling parenting and child-perceived mother-child attachment independently in the associations.

For the indirect paths through maternal controlling parenting in the associations between maternal ACEs and adolescent behavioural difficulties, the current results were consistent with a previous longitudinal study of preadolescents reporting a mediating effect of controlling parenting between maternal abuse history and preadolescent self-regulation difficulties (Delker et al., 2014), suggesting controlling parenting could be a substantial mediator between maternal ACEs and mental health outcomes among preadolescents and adolescents. Both this and our own findings suggest that maternal childhood adversity exposures are significantly and positively associated with maternal controlling parenting. This is a novel finding, albeit consistent with previous longitudinal work on intergenerational transmission of parenting, which identified that grandmothers but not grandfathers parenting behaviours were intergenerationally associated with parenting behaviours for both parents (Madden et al., 2015).

Alongside links to attachment and social learning approaches, this is also consistent with predictions from self-determination theory, suggesting controlling parenting behaviours that are inappropriate to the development stage may be perceived by the adolescent as over-controlling, invasive, and disregarding individual autonomy, impacting on self-regulation and mental health (Ryan & Deci, 2017; Soenens & Vansteenkiste, 2010, 2020).

With regard to the indirect effect through child-perceived mother-child attachment, our results differ from previous findings (Thomas-Giyer & Keesler, 2021). However, mother-child attachment was measured differently in the two studies, whereas Thomas-Giyer and Keesler (2021) measured attachment through mothers’ self-report the current study measured through children’s self-report (i.e., child-perceived). Further, families in Thomas-Giyer and Keesler’s study were participating in a family wellbeing promotion programme, in which social support could moderate (i.e., buffer) the relationship between maternal ACEs and child mental health difficulties and further lead to a biased conclusion of null effect (Thomas et al., 2018; Racine et al., 2018a, 2018b).

Maternal controlling parenting was also negatively associated with child-perceived mother-child attachment and child-perceived mother-child attachment further negatively associated with adverse adolescent mental health outcomes, consistent with findings for mechanisms implicated child externalizing behaviours in children aged 4 to 6 years old (Roskam et al., 2011).

Serial mediation indicated that maternal controlling parenting and child-perceived mother-child attachment completely mediated the association between maternal ACEs and adolescent externalizing behaviours, and partially mediated the associations between maternal ACEs and adolescent internalizing behaviours and anxiety. The partial mediation effect for the latter finding suggests additional pathways may exist between Maternal ACEs and adolescent internalizing behaviours. For instance, maternal depression risk still significantly predicts adolescent internalizing behaviours even after controlling for maternal ACEs, maternal controlling parenting, and child-perceived mother-child attachment. This is consistent with fndings regarding the effect of maternal exposures to prenatal and postnatal depression on the association between maternal ACEs and adolescent internalizing and externalizing behaviours (Letourneau et al., 2019).

Regarding the association between maternal ACEs and adolescent anxiety, our results expand the previous literature. suggesting a possible mechanism between maternal ACEs and adolescent anxiety, whilst not simply subsuming anxiety into emotional difficulties (Haynes et al., 2020) or internalizing behaviours (Cooke et al., 2019; Khoury et al., 2022; Shih et al., 2023). Further, few studies (Wang et al., 2022) have examined the association between maternal ACEs and child anxiety.

Our findings suggest a more complex model, rather than a causal mechanism from maternal health through to adolescent outcomes, which could be perceived as mother blaming. This is consistent with Bronfenbrenner’s bioecological model emphasizing that each level of influence impacts upon the ecological system and the developing child may interact with each other, and the influences between these levels and factors are bidirectional (Bronfenbrenner, 1977, 2001). Although maternal ACEs may influence child mental health outcomes through maternal controlling parenting and child-perceived mother-child attachment, child mental health difficulties may also increase parenting stress and may lead to negative parenting behaviours and deterioration of child-perceived mother-child attachment and mental health outcomes (Fang et al., 2022; Mak et al., 2020). Meanwhile, other environmental factors in the ecological system may also affect the mental health outcomes of young people (Kieling et al., 2011). A holistic model that integrates maternal, adolescents’, and other environmental factors into comprehensive consideration may foster a better understanding of intergenerational transmission mechanisms in youth mental health.

## Limitations

We note several limitations. First, maternal ACEs were accessed retrospectively and are thus subject to recall and motivation bias (Henry et al., 1994). However, a few literatures suggest retrospective ACEs reporting has sufficient validity for research as people usually underestimate their ACEs exposures rather than overestimating, which may result in a conservative estimate on the impact of maternal ACEs (Ferusson et al., 2000; Hardt & Rutter, 2004). Second, controlling parenting assessment was self-reported by mothers, and the results may be influenced by recall and social desirability bias (Bornstein et al., 2015; Morsbach & Prinz, 2006). Third, difficulties, measured by the SDQ scale, were also self-reported by cohort children, which may also be influenced by social desirability bias. In the meantime, although self-report SDQ has been well validated previously (e.g., Goodman, 2001; Goodman et al., 2010; Van Roy et al., 2008), the externalizing behaviour measurement was slightly under the ideal internal consistency cut-off (Nunnally, 1978). Finally, this study was conducted under a cross-sectional research design, which may have limited use for inferring causal relationships; therefore, a further longitudinal study is needed to confirm the findings from this study (e.g., Barnett et al., 2023; Savitz & Wellenius, 2023).

## Implications and Conclusions

Despite the above limitations, this study found a consistent and significant mediating effect via maternal controlling parenting and child-perceived mother-child attachment in the association between maternal ACEs and adolescent behavioural difficulties, both of which are possible intervention points in clinical practice. Clinical routine screenings of maternal ACEs history, risk factors for maternal controlling parenting, and child-perceived mother-child attachment may provide healthcare and child welfare services better opportunities for early prevention, identification, and intervention on parental ACEs, parenting traits, mother-child attachment, and child mental health difficulties, and all these clinical measures could help to interrupt the intergenerational cycle of childhood adversity (Koita et al., 2018; Shih et al., 2023). This study also emphasizes the importance of attending to both maternal and children’s factors in relation to intergenerational transmission of risk. Future studies should further examine this effect in longitudinal studies to confirm these findings. In the meantime, future studies should also examine the effect of paternal ACEs to understand better the intergenerational associations between parental ACEs and children’s mental health outcomes. Future research could also explore potential buffers and intervention strategies for young people as well as adults who had a child adversity history to cope and thrive with these adversities, including resilience and social support (Fritz et al., 2018; Thomas et al., 2018).

## Data availability

Due to Growing Up in Scotland access licensing arrangements the authors do not have permission to share data.

## Acknowledgement

The authors thank all participating children and their parents/caregivers in the Growing up in Scotland birth cohort 1.

